# Lifelong association of disorders related to military trauma with subsequent Parkinson’s disease

**DOI:** 10.1101/2021.08.21.21262374

**Authors:** Gregory D. Scott, Lee E. Neilson, Randy Woltjer, Joseph F. Quinn, Miranda M. Lim

## Abstract

**Background:** Trauma-related disorders such as traumatic brain injury (TBI) and posttraumatic stress disorder (PTSD) are emerging as risk factors for Parkinson’s Disease (PD), but their association with development of PD and independence from comorbid disorders remains unknown. This case-control study examines TBI and PTSD related to early trauma in military veterans.

**Methods:** PD was identified by ICD code, recurrent PD-specific prescriptions, and availability of 5+ years of earlier records. Validation was performed by chart review by a movement disorder-trained neurologist. Controls were matched 4:1 by age, duration of preceding health care, race, ethnicity, birth year, and sex. TBI and PTSD were identified by ICD code and onset based on active duty. Association and interaction were measured for TBI and PTSD with PD going back sixty years. Interaction was measured for comorbid disorders.

**Results:** 71,933 cases and 287,732 controls were identified. TBI and PTSD increased odds of subsequent PD at all preceding five-year intervals back to year -60 (OR range: 1.5 [1.4,1.7] to 2.1 [2.0,2.1]). TBI and PTSD showed synergism (synergy index range: 1.14 [1.09,1.29] to 1.28 [1.09,1.51]) and additive association (OR range: 2.2 [1.6,2.8] to 2.7 [2.5,2.8]). Chronic pain and migraine showed greatest synergy with PTSD and TBI. Effect sizes for trauma-related disorders were comparable to established prodromal disorders.

**Discussion:** TBI and PTSD are associated with later PD and are synergistic with chronic pain and migraine. These findings provide evidence for TBI and PTSD as risk factors preceding PD by decades and could aid in prognostic calculation and earlier intervention.

## INTRODUCTION

Identification of risk factors for Parkinson’s disease (PD) is important for understanding the pathogenesis, for prognostic calculation, and for earlier diagnosis of PD. One category of risk factors emerging in recent literature relates to war-related illness such as traumatic brain injury (TBI) and post-traumatic stress disorder (PTSD). Both disorders have been recently implicated as PD risk factors, but their association with development of PD, timing relative to PD onset, and independence from other risk factors remains unknown.

TBI is the better studied of these two risk factors, and a subset of human studies have implicated TBI as an acquired PD risk factor^1,2^. Mechanistic evidence in animal model systems and humans have supported this link by showing accumulation of pathologic proteins including α-synuclein after brain injury^3,4^. Human studies have also linked brain injury related to repetitive head injury to Lewy body disease^5^. However, some studies have suggested a reverse causation model where TBIs are the result from undiagnosed early PD^6^ (e.g. due to postural instability). Although there is still lingering debate, recent meta-analyses and those studies with a longer lag time support TBI to be a true risk factor for PD^1,2^. Moreover, association studies suggest TBI’s impact on PD risk is far greater in those who sustain early-in-life TBI, considered before the age of 30^7^. These data highlight the importance of TBI as a PD risk factor but also the need for increased study duration and details on timing in order to disentangle associations. The lifelong timecourse of TBI-conferred risk and its interaction with comorbid disorders remains unknown.

The link between PTSD and PD has only recently been identified. Mechanistically, animal studies have linked chronic stress to aggregation of toxic proteins (e.g. synuclein) and neurodegeneration ^8^. Epidemiological studies by our group and others have found an association between PTSD and RBD ^9,10^ and two larger studies (including one prospective study) reported direct associations between PTSD and PD^11,12^. However, the reported associations of PTSD with PD were not controlled for important confounders (e.g. smoking) and had shorter lag times susceptible to potential reverse causation.

Interactions between trauma-related disorders are unknown and could potentially cause more-than-additive (i.e. synergistic) effects on increasing PD risk. This is particularly important to study for co-occurring disorders with similar causes that could confer any manner of positive or negative impact on PD risk. TBI and PTSD are two such disorders that often co-exist as hallmarks of chronic neurobehavioral symptomatology along with other comorbid sleep, pain, mood/anxiety, and cognitive disturbances. As some of these comorbid disorders are potentially treatable, it is important to define their interaction with TBI, PTSD, and later PD.

Overall, we hypothesized that prevalence of TBI and PTSD would be greater in individuals that later developed PD, compared to matched-control cases who did not develop PD. Furthermore, we hypothesized that TBI and PTSD would interact in a synergistic manner to increase PD risk. In order to address these questions, we performed a retrospective case-control study within the United States Veterans Health Administration, a population enriched for early trauma, TBI, PTSD, and PD.^13^

## METHODS

### Research design, data source, and institutional approval

A retrospective case-control study design was used to measure the association of TBI and PTSD with later development of PD. Data was obtained from the Veterans Health Administration Corporate Data Warehouse (CDW) under IRB approval (MIRB #04744) using a waiver of participant consent. All time periods available within the CDW were included in the study. Dated ICD codes in the CDW began in 1999 as ICD digitization was implemented at VA hospitals between 1999-2005.

### Case definition and validation

PD was defined by: 1) age greater than 40 years old, 2) the presence of one or more PD ICD-9 (332.0) or ICD-10 (G20) codes, 3) having at least 5 years of ICD-coded medical records before the first PD ICD code, and 4) having two PD medication prescriptions filled. PD medication was defined by the VA nationwide common data model for operations and research (OMOP)^14^ (**eTable 1**). In order to increase specificity, we created a secondary definition. In addition to meeting all previous criteria, these cases required affirmative mention of PD in at least two VA neurology notes, separated by a minimum of one year in time, and starting at least one year after the first PD ICD code. Thus, each case would have a minimum 2 years of free-text records referring to PD as a chief complaint. Regular expressions and iterative training were used to eliminate negations and ambiguous statements. Both definitions were validated against manual chart review as a gold standard in 150 randomly selected cases. The index date was defined as the date of the first PD ICD code in cases.

### Control selection

Four controls were chosen for each PD case by randomized selection and matching by sex, ethnicity, race, smoking status^15^, and birth year. Controls were also selected for those with at least the same number of years of historic medical records before the index date as their matched case. The index date for controls was the same age (month and year) as the time of first coded PD diagnosis in their matched case^16,17^. 21,882,307 Veterans were potential controls at the time of writing this manuscript.

### Variables definition

Extraction of race, ethnicity, sex, and birth year was from their own distinct fields in the CDW (see eMethods for field names) and smoking information was categorized using a crosswalk method validated previously at the VA^15(p20)^. Missing data was coded as unknown rather than performing imputation and assuming missingness was random.

Comorbid disorders were defined by diagnostic codes (ICD-9, ICD-10) that were collected for each participant within twenty years of the index date. ICD codes for each disorder category are listed in **eTable 2** in the Supplement. For TBI and PTSD in this military Veteran cohort, person-specific onset was defined as the midpoint of active duty. Only TBI and PTSD with onset 5+ years before the index date is included in order to mitigate reverse causation. We do not use dates of ICD codes because VA records begin years-to-decades after active duty and would severely underestimate onset time of trauma exposures. To parallel PD case ascertainment, a secondary definition for TBI and PTSD was created to improve specificity, further enrich for true military-related TBI and PTSD, and further eliminate cases caused by non-military factors in the Veteran population. This required that these diagnoses are “service connected”, i.e. explicitly indicated in the record to be connected to the participants’ active military service. We suspect usage and data completeness of the “service connected” field was low. For other disorders, the first instance of a relevant ICD code defined its onset. The possibility of multiple distinct episodes of the same disorder is not captured since the majority of repeated codes are related to the initial diagnosis^18^.

### Statistical Analyses

For TBI, PTSD, and other clinical disorders defined by ICD codes, yearly prevalence was calculated before the index date. Association of each disorder with PD was calculated using conditional logistic regression adjusted for race, sex, smoking status, ethnicity, and birth year. Odds ratios (OR) and 95% confidence intervals (CI) are shown in figures at 5-year and yearly epochs. P values were adjusted for multiple comparisons using Bonferroni correction and significant results were defined as p-value less than 5 × 10^−2^. Analysis was conducted using RStudio (RStudio Team (2021). RStudio: Integrated Development for R. RStudio, PBC, Boston, MA URL http://www.rstudio.com/).

### Interaction analysis

Interaction was measured using Synergy Index (SI), a recommended metric for case controls studies^19^. Indices and confidence intervals were quantified using the interactionR package in R studio^20^. SI greater than 1 indicates a positive interaction or greater than additivity.

### Data Sharing

The United States Veterans Affairs legally restricts access to data which includes sensitive and identifying patient information. The data used for this study are not permitted to leave the Veterans Affairs firewall without a Data Use Agreement. Behind the firewall, data are made available with an approved study protocol.

## RESULTS

### Cohort Characteristics

71,933 patients fulfilled the primary PD case definition and 287,732 controls (matched 4:1) were identified (**Figure 1**, left side). The cohort was predominantly male (98.4%), white (81.4%), non-Hispanic (88.3%), and elderly (mean age at index date 74.8 ± 8.9 years old) (**Table 1**). For smoking status, the cohorts’ proportions were split between former smokers (39.9%) and never smokers (37.1%) with a lower proportion of current smokers (20%) and unknown (2.9%). Manual chart review was performed for 150 randomly selected (**Figure 1**, right side), and 118 true positives were identified for a positive predictive value of 78.6%. The most common specific types of false positives were drug-induced parkinsonism (n=7), and essential tremor (n=4).

**Figure 1:**
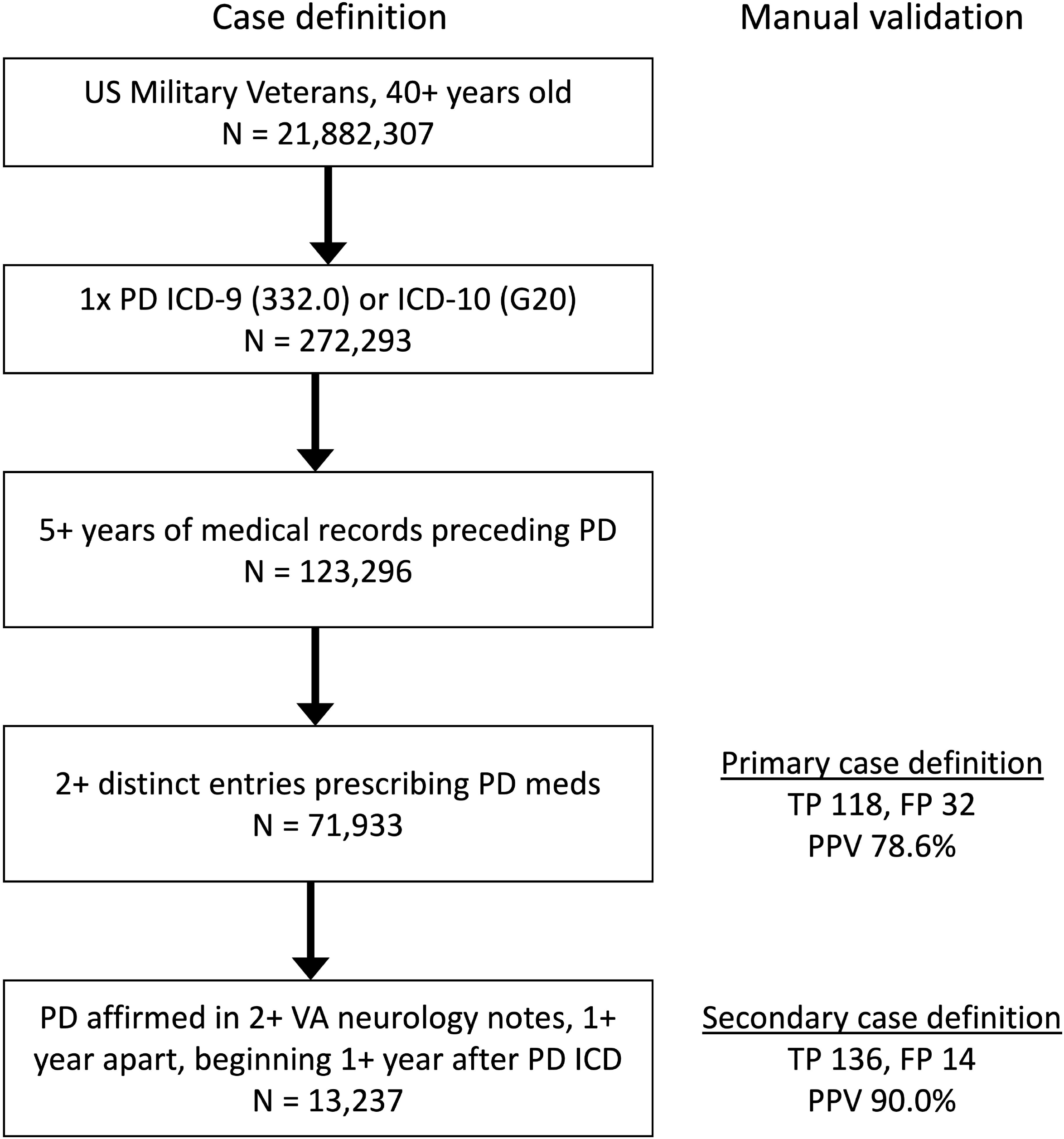
PD case definitions. Flowchart (left) shows stepwise criteria and case counts for primary and secondary PD case definitions. Definitions were validated (right) by manual review of medical records. TP = true positive, FP = false positive, PPV = positive predictive value.

**Table 1:**
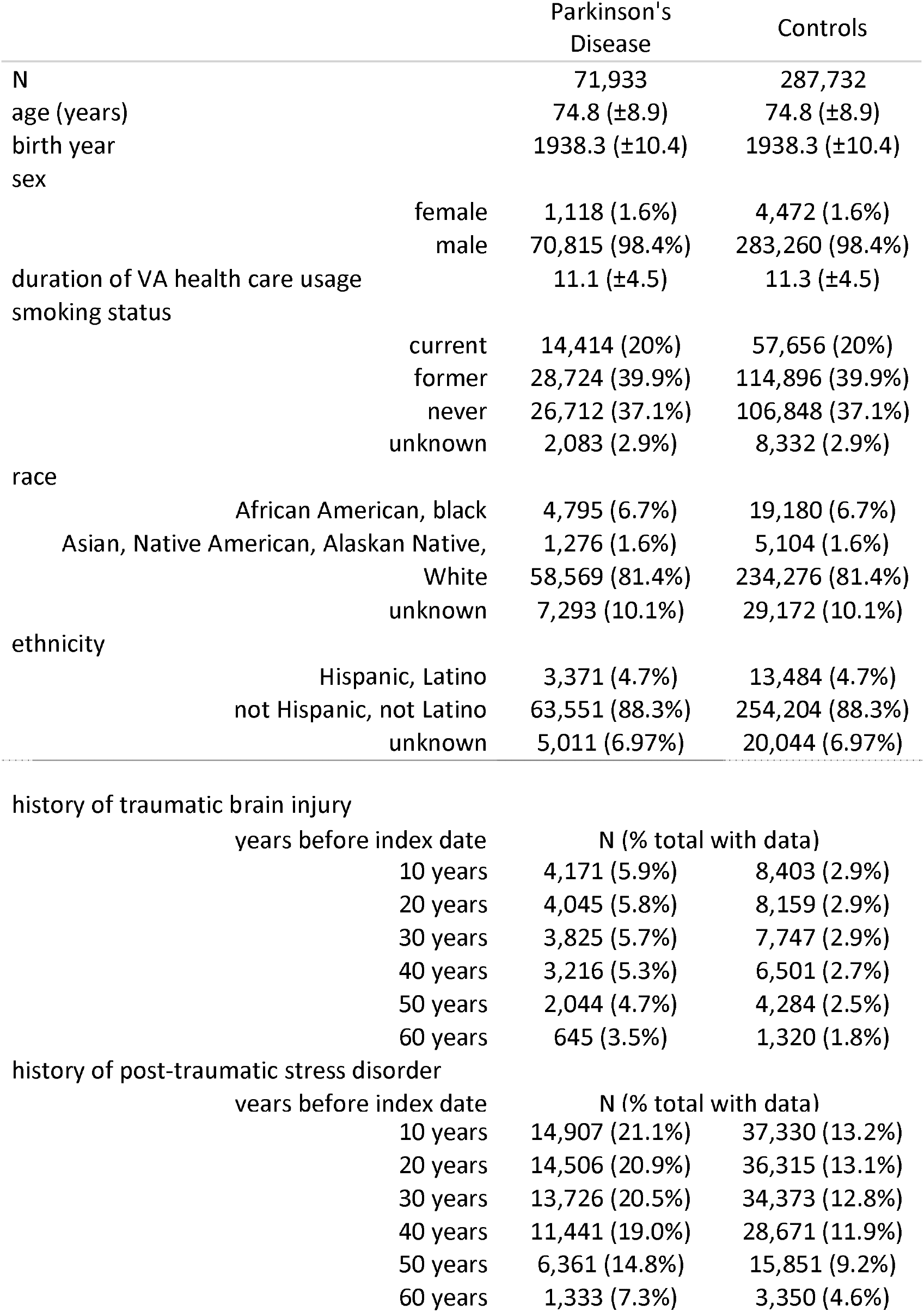
Cohort characteristics used for matching

In an attempt to improve specificity for secondary analyses, we next identified 13,237 Veterans fulfilling the secondary PD definition, requiring affirmative mention of PD in at least two VA neurology notes. Validation by chart review identified 136 true positives and 14 false positives, yielding an improved positive predictive value of 90.0%.

### Traumatic brain injury and post-traumatic stress disorder

Prevalence of TBI and PTSD was calculated at 5-year time epochs across a 60-year duration before the index date of PD diagnosis (**Figure 2A-B**). Raw numbers of cases and controls with TBI and PTSD are also shown at ten-year epochs in **Figure 2A-B** and **Table 1**. TBI had an average prevalence of 2.6% ± 0.3% in controls and 5.3% ± 0.7% in cases who were later diagnosed with PD. PTSD prevalence was on average 11% ± 2.9% in controls and 17% ± 4.7% in cases. Association analysis was performed and both TBI and PTSD had significantly increased odds of PD at each time epoch extending back 60 years. The association between TBI and PD was calculated at 5-year time epochs and ranged from OR = 2.0 [1.8,2.1] to 2.1 [2.0,2.1]. The association between PTSD and PD across the same time period ranged from OR = 1.5 [1.4, 1.7] to 1.9 [1.9, 2.0].

**Figure 2:**
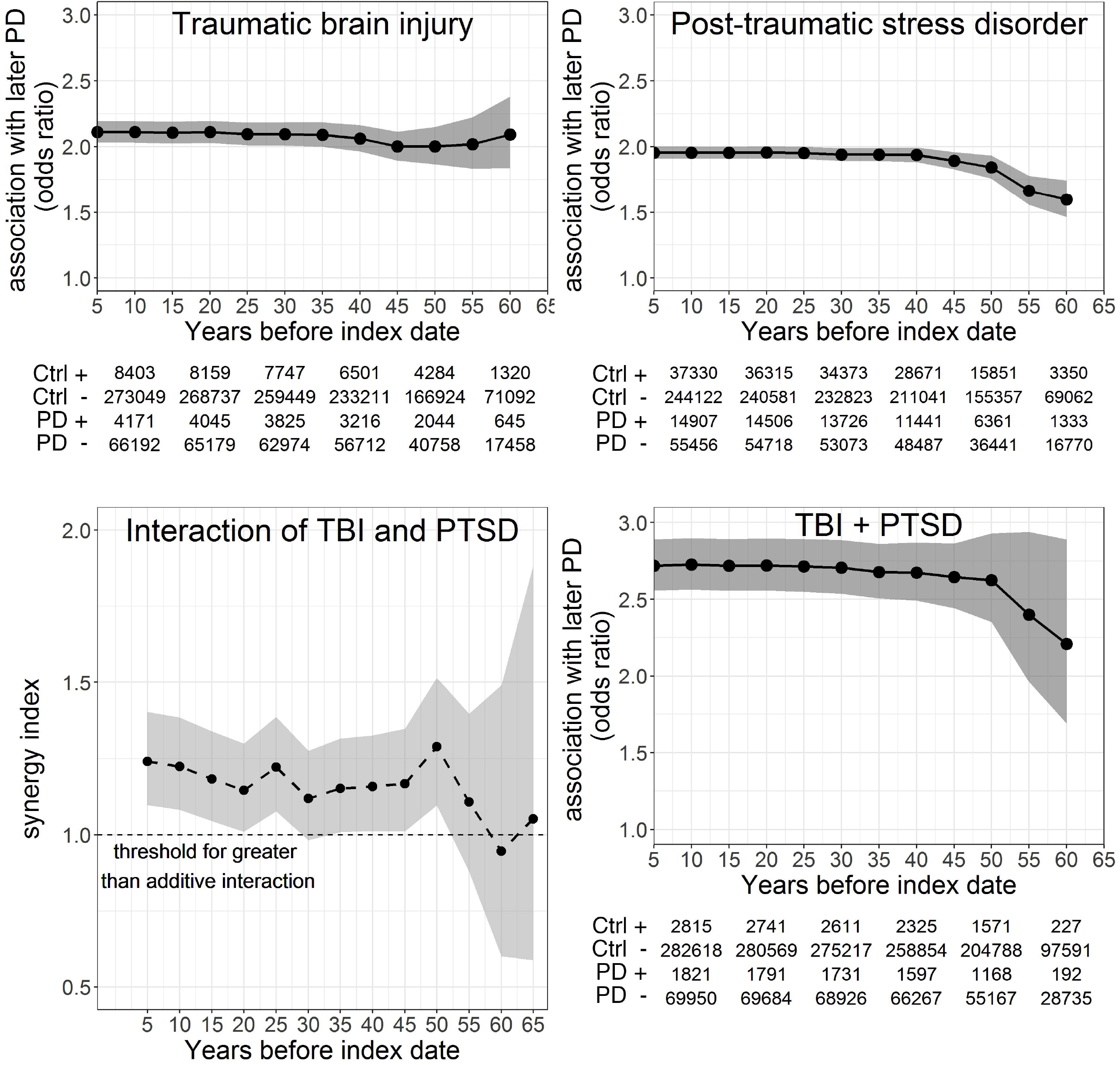
Association of trauma-related disorders with Parkinson’s disease (PD). Panel A: Association of traumatic brain injury (TBI) with subsequent PD. Panel B: Association of Post-traumatic stress disorder (PTSD) with subsequent PD. Bottom panels: Interaction of TBI and PTSD in association with subsequent PD. Panel C: Interaction of TBI and PTSD in influencing odds of subsequent PD. Panel D: Additive association of TBI plus PTSD on later PD diagnosis. For association analyses, odds ratios (black circles) and 95% confidence intervals (gray) shown at five-year intervals beginning at midpoint of active military duty and extending up to 5 years before the index date (year 0, date of PD diagnosis). Tables show number of cases and controls with (+) and without (-) trauma-related disorders. For interaction analysis, synergy index (small black circles) and 95% confidence intervals (light gray) shown at five-year intervals beginning at midpoint of active military duty and extending up to 5 years before the index date.

Analysis was repeated using the secondary PD definition (**eFigure 1**), with similar patterns emerging: mean TBI prevalence was 2.4% ± 0.3% in controls and 4.3% ± 0.4% in cases; mean PTSD prevalence was 13% ± 4.2% in controls and 22% ± 0.6% in cases. Moreover, association analyses revealed significantly increased odds of PD at each epoch extending back 60 years for both TBI (range of OR = 1.7 [1.6, 1.9] to OR = 2.4 [1.5,3.9]) and PTSD (range of OR = 1.9 [1.9, 2.0] to OR = 2.1 [1.7, 2.6]).

Analysis was again repeated on the primary PD definition after limiting TBI and PTSD diagnoses to those where a “service connected” field had been checked (**eFigure 2**), in order to improve specificity of military-related trauma diagnoses. Notably, case counts and prevalence for TBI dropped two orders of magnitude. Nevertheless, the same significant pattern emerged:

TBI had an average prevalence of 0.05% ± 0.01% in controls compared to 0.09% ± 0.03% in cases, and PTSD prevalence was 4.5% ± 1.2% in controls versus 7.5% ± 2.0% in cases. Significant association for TBI and PD began at year -45 and ranged from OR = 1.5 [1.03, 2.1] to OR = 2.0 [1.5, 2.6]. The association between PTSD and PD was significant across 60 years and ranged from OR = 1.7 [1.5, 1.9] to OR = 1.8 [1.7, 1.8].

### Interaction and combined PD association of TBI and PTSD

Since TBI and PTSD are commonly co-occurring conditions, we next aimed to explore whether their combination influenced PD risk beyond additivity. Interaction analysis was performed between TBI and PTSD (**Figure 2C**). Synergy index (SI) and 95% confidence intervals were measured at 5-year intervals across 60 years before PD diagnosis. SI values were significantly above additivity up to year -50 at all but one time epoch. Values significantly above additivity ranged from SI = 1.14 [1.09, 1.29] to SI = 1.28 [1.09, 1.51]. The one epoch which did not reach statistical significance was at year -30 with SI = 1.11 [0.98, 1.27].

Prevalence of individuals with both TBI and PTSD was calculated at 5-year time epochs across a 60-year duration before the index date / PD diagnosis (**Figure 2D**). Raw numbers of cases and controls with both TBI and PTSD are shown at ten-year epochs in tables of **Figure 2D**. TBI+PTSD had a greater numerical difference between PD and controls when combined versus when compared to either disorder separately. TBI+PTSD had an average prevalence of 0.8% ± 0.2% in controls and 2.1% ± 0.5% in cases who were later diagnosed with PD. Association analysis was performed and having both TBI and PTSD significantly increased odds of subsequent PD at each time epoch extending back 60 years. This association was calculated at 5-year time epochs and ranged from OR = 2.2 [1.6, 2.8] to OR = 2.7 [2.5, 2.8].

### Interaction of TBI and PTSD with other disorders

Since TBI and PTSD are comorbid with other pain, sleep, cognitive, mood, and prodromal disorders, we performed interaction analysis between combinations of TBI and PTSD with these disorders (**Table 2**). Synergy index (SI) was calculated at one-year time intervals across ten years from year -1 to year -11. The disorders with the largest significant positive interaction for both PTSD and TBI were migraine with average SI = 1.61 ± 0.33, chronic pain with average SI = 1.55 ± 0.34, and sleep apnea with average SI = 1.40 ± 0.19.

**Table 2:** Disorders synergistic with PTSD and TBI in increasing odds of later PD

|  | Synergy Index |
| --- | --- |
|  | Mean (SD) |
| Positive interaction with both PTSD and TBI |  |
| Migraine | 1.61 ($\pm 0.33$) |
| Chronic pain | 1.55 ($\pm 0.34$) |
| Sleep apnea | 1.40 ($\pm 0.19$) |
| Positive interaction with just PTSD |  |
| Cognitive impairment | 1.55 ($\pm 0.07$) |
| Constipation | 1.47 ($\pm 0.54$) |
| Orthostatic hypotension | 1.47 ($\pm 0.42$) |
| Positive interaction with just TBI |  |
| Depression | 1.46 ($\pm 0.05$) |
| Anxiety disorder | 1.34 ($\pm 0.15$) |

Greater than additivity is indicated by values above 1 for Synergy Index. Values shown are significant, $p < 0.05$ .

PD association analysis was performed for individuals with TBI, PTSD, and either chronic pain or migraine (**Figure 3**). The yearly prevalence for participants with chronic pain, TBI, and PTSD was on average 0.14% ± 0.08% in controls and 0.45% ± 0.24% in cases and PD association ranged from OR = 2.9 [2.6, 3.4] to OR = 3.3 [2.2, 5.0]. The yearly prevalence of migraine, TBI, and PTSD was on average 0.14% ± 0.07% in controls and 0.40% ± 0.03% in cases and PD association ranged from OR = 2.7 [2.1, 3.4] to OR = 3.1 [2.7, 3.6]. Results were compared to International Parkinson and Movement Disorder Society (MDS) prodromal disorders (**Figure 3**).

**Figure 3:**
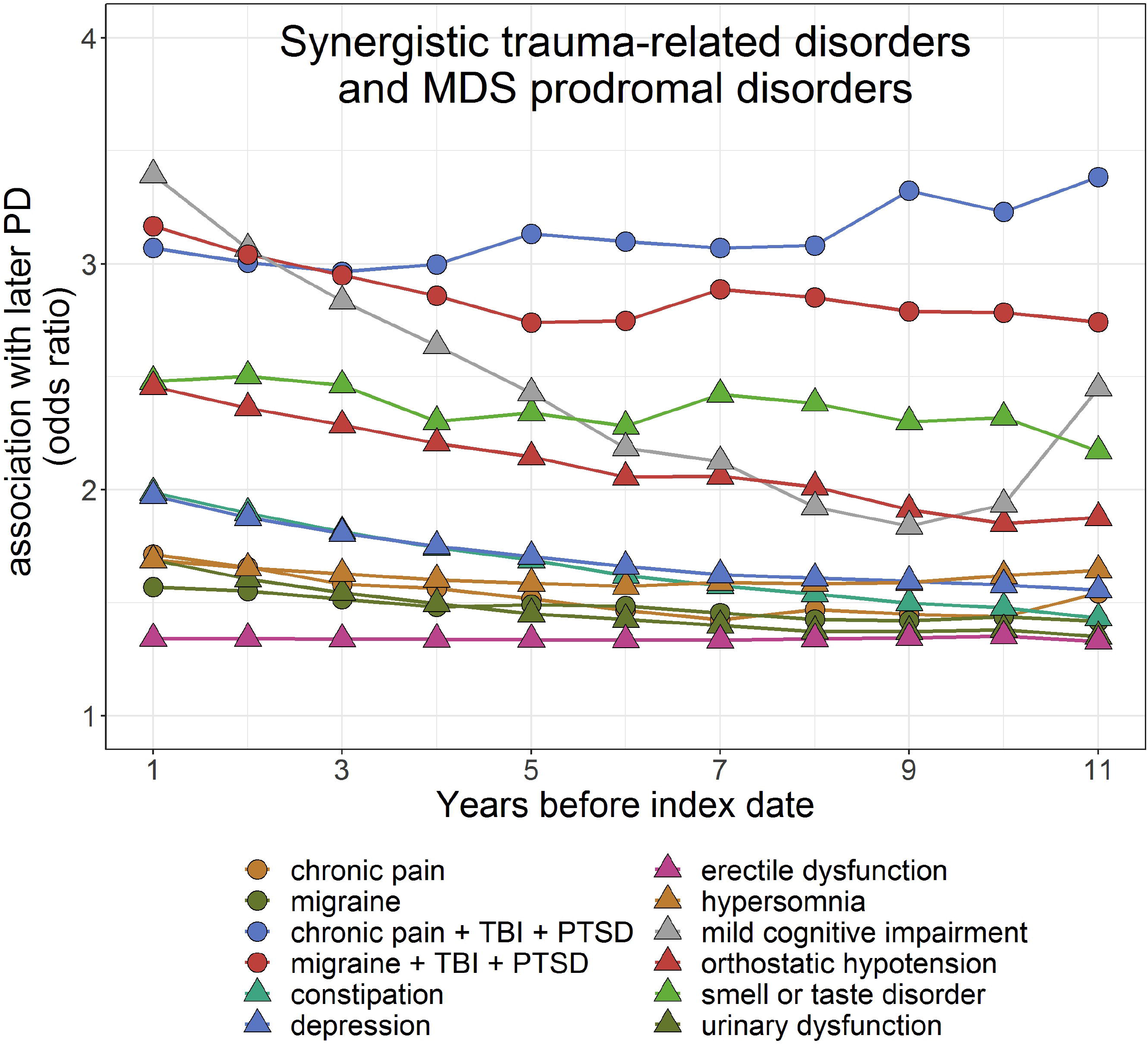
Synergistic disorders related to military trauma and also MDS prodromal disorders association with PD. Significant odds ratios (p < 0.05, Bonferroni’s correction) are shown for trauma-related disorders (circles) and MDS prodromal disorders (triangles) at one-year intervals beginning at the first appearance of the relevant ICD code and extending up to 1 year before the index date (year 0, date of PD diagnosis). REM-behavior sleep disorder is not shown as it demonstrated a larger effect size ranging from OR = 5.4 [3.0, 9.8] to OR = 8.5 [7.3, 9.9].

## DISCUSSION

We leveraged a nationwide electronic record of patients enriched for military trauma and show that TBI and PTSD are associated with later PD. Strengths of this study include improving the specificity of PD diagnosis, utilizing active duty timing information for early-in-life neurotrauma, and testing comorbid disorder combinations. We also showed that TBI and PTSD are synergistic with chronic pain and migraine and that synergy of TBI/PTSD with chronic pain and migraine further increased odds of PD. These findings provide evidence for TBI and PTSD as risk factors preceding PD by decades, and could aid in prognostic calculation, earlier diagnosis, and earlier intervention for PD.

### Association between TBI and PD

Data confirm TBI as a significant early-in-life risk factor for PD. Of note, the average age during active duty is 28-years-old^21^. The current effect sizes and ages match those of a smaller non-military study that reported an effect size of 2.3 (confidence intervals not available) in those reporting head injury in their teens-to-twenties approximately 40-50 years before development of PD^7^. The effect size is also similar to shorter duration studies of VA participants (1.7 [1.5,1.9]) ^2^ and slightly greater than pooled results of 15 other studies with shorter lag times (1.4 [1.1,1.7]) ^7,22,23^. In addition to extending upon human epidemiologic data, the current study corroborates chronic pathologies reported by animal studies of neurotrauma^24–28^. These studies have catalogued long-term phenomena including progressive dopaminergic cell loss, decreased tyrosine hydroxylase expression and increased alpha-synuclein accumulation in surviving dopaminergic neurons. Interestingly, these studies also found non-neuronal and acellular aberrancies including increased microglial inflammation, and dysregulation of soluble alpha-synuclein, in some cases persisting into the chronic phase of recovery. Additional studies are needed to search in human tissues for pathologies of lifelong trauma-related PD risk.

### Association between PTSD and PD

Results also show PTSD is associated with increased odds of subsequent PD and is not just a surrogate for TBI-associated PD risk. Our data are similar to the association between PTSD and PD previously reported in a large retrospective study^11^ and multiple other studies, within and outside the VA ^2,11,12,29^. Mechanistically, animal studies have found a link between chronic stress, aggregation of toxic proteins (e.g. synuclein), and neurodegeneration ^8^. Given these findings, translational studies of PTSD pathology in human biopsy and autopsy tissues are needed to pair with longitudinal clinical data.

### Interaction between TBI, PTSD and PD

The combination of TBI and PTSD further increased odds of PD beyond either factor alone. These results are not altogether surprising, as we and others have shown that the combination of TBI and PTSD in Veterans synergizes to produce higher neuropsychiatric symptom burden^10,30,31^, more profound disability^31^, and higher rates of REM sleep behavior disorder (RBD)^10^. Potential mechanisms underlying the additive effects of comorbid TBI and PTSD are still unclear, but animal models suggest that neurodegeneration is accelerated by chronic stress and sleep disruption, two disorders increased in TBI and PTSD^1^. Importantly, these data do not support the view that PTSD and TBI confer redundant risk and are not worth considering independently in PD subtyping and neuroprotection efforts.

### Synergy between TBI, PTSD and other comorbid disorders

A novel approach in this study was to explicitly examine comorbid disorders and screen for high-yield interactions. Interestingly, greater than additive association of developing PD in those with TBI and PTSD was strongest for chronic pain and migraine. The triad of TBI, PTSD, and chronic pain has been termed the “Polytrauma Clinical Triad” – a syndrome recognized for its association with disability and poor outcomes^31^. Our results suggest that Veterans with the Polytrauma Clinical Triad may also be at higher risk for neurodegenerative disorders. Additional human studies with more detailed phenotyping of pain and migraine will help identify what makes these potent mixtures with TBI and PTSD.

### Limitations

Limitations of this study include those associated with retrospective case-control studies reliant on diagnostic coding. In general, using ICD codes for other disorders typically represents a “possible” or “probable” level of certainty depending on usage and classification criteria. Previous studies measuring the test performance of ICD codes at the VA report 75.6% PPV for PD at a single VA movement disorder specialty center^18^, 82% PPV and 84% sensitivity for PTSD across the VA^34^, and 84.6% PPV for TBI at the VA but with low sensitivity of 55.4%^35^. Note that the PPV result for PD ICD codes is inflated by restricting to one specialty center, and in studies using all medical providers, PPV is 46% and sensitivity is 80%^36^. Our PD case definitions were applied across the entire VA system and not just a specialty center yet yielded higher positive predictive values of 78.6% and 90.0%, depending on our case definition. We suspect miscoding would only have lowered the observed effect sizes. For example, miscoding PD as drug-induced parkinsonism, the most common false positive in our study, is less likely to have the same pre-diagnosis disorders ten years earlier. Capturing additional certainty in the current study would have required a prohibitive amount of extraction of non-standardized data from terabytes of free-text medical notes. We also suspect ICD insensitivity underlies low case ascertainment of entities like REM behavior sleep disorder in the current study. The prevalence of RBD in the current study is far lower than previous studies in both PD and control groups and we suspect underpowers interaction analysis with PTSD and TBI. A final limitation of ICD codes is misdiagnosis which can skew results. Misdiagnosis will be mitigated in future studies by newer tools such as the validated questionnaire, the Comprehensive TBI Evaluation program, CTBIE^2^).

An additional limitation includes the use of birth year as an adjustment variable, which could be skewed towards Vietnam-era Veterans who may have had exposure to Agent Orange. While officially recognized by the VA in service connection to PD, at the time of this writing, Agent Orange has still not been proven as an established risk factor for PD and therefore was not included as an adjustment variable.

Another important limitation for generalizing findings is under-representation of non-white races, Hispanic ethnicity, and female sex. Under-representation of female sex, for example, may skew disorders that covary by sex such as PTSD, depression, anxiety, headache, migraine ^37–40^. Under-representation of race/ethnicity may falsely inflate sensitivity of diagnoses like TBI where VA studies have shown under-diagnosis amongst Black, Hispanic, and Asian service members compared to Whites^41^.

A final limitation of a retrospective case-control design is disentangling the PD risk contribution of two co-occurring disorders. It is suggestive, albeit inconclusive, when two disorders (e.g. PTSD and cognitive impairment) have synergistic effects on developing PD. The current study cannot resolve independent risk and relies on studies with prospective design and more detailed patient-specific onset timing to determine independent risk of say PTSD versus cognitive impairment. Accurate risk estimation is one reason MDS research criteria require prospective studies when designing a prodromal risk calculator^42^. This limitation applies in general to other co-occurring diseases where, for example, TBI and PTSD and PD each may directly cause problems of sleep, cognition, pain, and anxiety/mood.

### Conclusions

Overall, these data found lifelong association of trauma-related disorders with subsequent PD. Interaction analysis uncovered synergism with comorbid chronic pain and migraine. These effects should help guide intervention trials and studies into distinct PD subtypes. We hope our results expand the search for putative acquired and preventable insults^43^ and jumpstart earlier prodromal screening tools^44^, which will be particularly impactful once effective neuroprotective therapy is available. Answering the questions of when and what disorders make PD more likely will be critical to understanding pathogenesis and spur innovative thinking about novel targets and optimal timing of potential interventions.

## Data Availability

Abstracted and fully de-identified data can be made available upon request with appropriate approvals.

## ACKNOWLEDGMENT

We would like to thank Priya Srikanth, MPH of the Biostatistics program at Oregon Health and Science University for biostatistics help. We would also like to thank the OHSU School of Medicine and Department of Pathology and the Parkinson’s Center of Oregon. This work was also supported in part by VA Biomedical Laboratory Research and Development Career Development Award Number 1 IK2 BX005760-01A1 to GS and VA Clinical Science Research and Development Merit Award I01 CX002022 to MML. All authors (G.D. Scott, L. E. Neilson, R. Woltjer, J.F. Quinn, M. M. Lim) report no disclosures relevant to the manuscript.

## AUTHOR ROLES

Design, writing, editing: GS, LN, ML, JQ, RW. Execution, analysis: GS, LN.

