## Supplement for "Lifelong association of disorders related to military trauma with subsequent Parkinson’s disease"

eMethods

Searches were performed using structured query language (SQL) within Microsoft SQL Server Management Studio (Microsoft Corporation, Redmond, WA). The CDW fields containing demographics and birth dates are named “PatSub_PatientRace”, “PatSub_PatientEthnicity”, “SPatient_Gender”, “SPatient_BirthDateTime”. The CDW fields containing ICD codes are named “Outpat_VDiagnosis”, “Outpat_WordloadVDiagnosis”, “Inpat_InpatientDiagnosis”, “Inpat_DischargeDiagnosis”, “Inpat_InpatientFeeDiagnosis”, “Inpat_PatientTransferDiagnosis”, “Inpat_PresentOnAdmission”, “Inpat_SpecialtyTransferDiagnosis”.

eTable 1: PD medication examples as defined by VA data warehouse “OMOP” common data model

| Carbidopa | Sinemet |
| --- | --- |
| Levodopa | Rotigotine |
| Entacapone | Zalapar |
| Pramipexole | Stalevo |
| Apomorphine | Istradefylline |
| Rasagiline | Azilect |
| Ropinirole | Comtan |
| Selegiline | Pergolide |
| Tolcapone | Eldepryl |
| (additional brand names for generic names listed above) | |

eTable 2

|  | ICD-9 or ICD-10 | Code |
| --- | --- | --- |
| Post-traumatic stress disorder |  |  |
| Post-traumatic stress disorder | ICD-10 | F43.1 |
| Post-traumatic stress disorder | ICD-9 | 309.81 |
| Prolonged Post-traumatic stress disorder | ICD-9 | 309.81 |
| Traumatic Brain Injury |  |  |
| Fracture of vault of skull | ICD-9 | 800.x |
| Fracture of base of skull | ICD-9 | 801.x |
| Other and unqualified skull fractures | ICD-9 | 803.x |
| Multiple fractures involving skull or face with other bones | ICD-9 | 804.x |
| Concussion | ICD-9 | 850.x |
| Cerebral laceration and contusion | ICD-9 | 851.x |
| Subarach. subdural and extradural hemorrhage folllowing injury | ICD-9 | 852.x |
| Other and unspecified intracranial hemorrhage following injury | ICD-9 | 853.x |
| Intracranial injury of other and unspecified nature | ICD-9 | 854.x |
| Intracranial injury | ICD-10 | S06.x |
| Concussion | ICD-10 | S06.0 |
| Traumatic Cerebral Edema | ICD-10 | S06.1 |
| Diffuse traumatic brain injury | ICD-10 | S06.2 |
| Focal traumatic brain injury | ICD-10 | S06.3 |
| Epidural hemorrhage | ICD-10 | S06.4 |
| Traumatic subdural hemorrhage | ICD-10 | S06.5 |
| Traumatic subarachnoid hemorrhage | ICD-10 | S06.6 |
| Other specified intracranial injuries | ICD-10 | S06.8 |
| Unspecified intracranial injury | ICD-10 | S06.9 |
| Traumatic brain compression and herniation | ICD-10 | S06.A |
| Fracture of vault of skull | ICD-10 | S02.0 |
| Fracture of base of skull | ICD-10 | S02.1 |
| Headache |  |  |
| Other headache syndromes | ICD-10 | G44 |
| Headache | ICD-10 | R51 |
| Periodic headache syndromes | ICD-10 | G43.C |
| Other headache syndromes | ICD-9 | 339 |
| Tension headache | ICD-9 | 307.81 |
| Headache | ICD-9 | 784 |
| Migraine |  |  |
| Migraine | ICD-10 | G43 |
| Migraine | ICD-9 | 346 |
| Chronic pain |  |  |
| Chronic bladder pain | ICD-10 | R39.82 |
| Chronic pain due to trauma | ICD-10 | G89.21 |
| Chronic pain syndrome | ICD-10 | G89.4 |
| Other chronic pain | ICD-10 | G89.29 |
| Chronic pain, not otherwise specified | ICD-9 | 338.29 |
| Chronic pain syndrome | ICD-9 | 338.4 |
| Chronic pain, postop, not otherwise specified | ICD-9 | 338.28 |
| Neck pain |  |  |
| Cervicalgia | ICD-10 | M54.2 |
| Cervicalgia | ICD-9 | 723.1 |
| Back Pain |  |  |
| Low back pain | ICD-10 | M54.5 |
| Backache, not otherwise specified | ICD-9 | 724.5 |
| Joint pain |  |  |
| Pain in limb, hand, foot, fingers, and toes | ICD-10 | M79.6 |
| Pain in joint | ICD-10 | M25.5 |
| Pain in joint | ICD-9 | 719.4 |
| Pain in limb | ICD-9 | 729.5 |
| Mild cognitive impairment |  |  |
| Mild cognitive impairment, so stated | ICD-10 | G31.84 |
| Mild cognitive impairment | ICD-9 | 331.83 |
| Cognitive Impairment |  |  |
| Age-related cognitive decline | ICD-10 | R41.81 |
| Oth symptoms and signs w cognitive functions and awareness | ICD-10 | R41.89 |
| Unsp symptoms and signs w cognitive functions and awareness | ICD-10 | R41.9 |
| Presenile delerium | ICD-9 | 290.11 |
| Presenile dementia | ICD-9 | 290.10 |
| Senile delerium | ICD-9 | 290.3 |
| Senile dementia | ICD-9 | 290.0 |
| Bipolar disorder |  |  |
| Bipolar disorder | ICD-10 | F31 |
| Bipolar disorder | ICD-9 | 296.0 |
|  |  | 296.4-7 |
|  |  | 296.80 |
|  |  | 296.89 |
| Anxiety disorder |  |  |
| Anxiety disorder due to known physiological condition | ICD-10 | F06.4 |
| Anxiety disorder, unspecified | ICD-10 | F41.6 |
| Generalized anxiety disorder | ICD-10 | F41.1 |
| Oth psychoactive substance dependence w anxiety disorder | ICD-10 | F19.280 |
| Other mixed anxiety disorders | ICD-10 | F41.3 |
| Other specified anxiety disorders | ICD-10 | F41.8 |
| Anxiety disorder associated with other diseases | ICD-9 | 293.84 |
| Anxeity state, not specified | ICD-9 | 300.0 |
| Generalized anxiety disorder | ICD-9 | 300.02 |
| Organic anxiety syndrome | ICD-9 | 293.84 |
| Depression |  |  |
| Adjustment disorder with depressed mood | ICD-10 | F43.2 |
| Adjustment disorder with mixed anxiety and depressed mood | ICD-10 | F43.23 |
| Major depressive disorder, recurrent | ICD-10 | F33 |
| Depressive episode | ICD-10 | F32 |
| Mood disord d/t physiol cond w major depressive-like epsd | ICD-10 | F06.32 |
| Mood disorder due to known physiol cond w depressv features | ICD-10 | F06.31 |
| Other depressive episodes | ICD-10 | F32.8 |
| Other recurrent depressive disorders | ICD-10 | F33.8 |
| Other specified depressive episodes | ICD-10 | F32.89 |
| Atypical depressive disorder | ICD-9 | 296.82 |
| Chronic depressive person | ICD-9 | 301.12 |
| Depressive disorder, not otherwise specified | ICD-9 | 311 |
| Major depressive disorder, recurrent, moderate | ICD-9 | 296.3 |
| Neurotic depressive state | ICD-9 | 300.4 |
| Presenile dementia with depressive features | ICD-9 | 290.13 |
| Adjustment disorder with depressed mood | ICD-9 | 309.0 |
| Prolonged depressive episode | ICD-9 | 309.1 |
| Adjustment disorder with mixed anxiety and depressed mood | ICD-9 | 309.28 |
| Senile dementia with depressive features | ICD-9 | 290.21 |
| Hypersomnia |  |  |
| Hypersomnia | ICD-10 | G47.1 |
| Hypersomnia not due to a substance or known physiol cond | ICD-10 | F51.1 |
| Hypersomnia with sleep apnea | ICD-9 | 780.53 |
| Hypersomnia, not specified | ICD-9 | 780.54 |
| Idiopathic hypersomnia with long sleep time | ICD-9 | 327.11 |
| Idiopathic hypersomnia without long sleep time | ICD-9 | 327.12 |
| Organic hypersomnia, not specified | ICD-9 | 327.19 |
| Recurrent hypersomnia | ICD-9 | 327.13 |
| Transient hypersomnia | ICD-9 | 307.44 |
| Circadian rhythm sleep disorder |  |  |
| Circadian rhythm sleep disorders | ICD-10 | G47.2 |
| Circadian rhythm sleep disorder | ICD-9 | 327.3 |
| Sleep apnea |  |  |
| Sleep apnea | ICD-10 | G47.3 |
| Sleep apnea, unspecified | ICD-9 | 780.5 |
| Sleep apnea, central/obstructive/organic | ICD-9 | 327.2 |
| REM behavior sleep disorder |  |  |
| REM sleep behavior disorder | ICD-10 | G47.52 |
| REM sleep behavior disorder | ICD-9 | 327.42 |
| Restless legs syndrome |  |  |
| Restless legs syndrome | ICD-10 | G25.81 |
| Restless legs syndrome | ICD-9 | 333.94 |
| Periodic limb movement disorder |  |  |
| Periodic limb movement disorder | ICD-10 | G47.61 |
| Periodic limb movement disorder | ICD-9 | 327.51 |
| Insomnia |  |  |
| Insomnia, unspecified | ICD-10 | G47.00 |
| Insomnia due to medical condition | ICD-10 | G47.01 |
| Adjustment insomnia | ICD-10 | F51.02 |
| Paradoxical insomnia | ICD-10 | F51.03 |
| Psychophysiologic insomnia | ICD-10 | F51.04 |
| Insomnia due to medical condition | ICD-10 | G47.01 |
| Oth insomnia not due to a substance or known physiol cond | ICD-10 | F51.09 |
| Insomnia due to other mental disorder | ICD-10 | F51.05 |
| Primary insomnia | ICD-10 | F51.01 |
| Insomnia due to mental disorder | ICD-9 | 327.02 |
| Insomnia with sleep apnea | ICD-9 | 780.51 |
| Insomnia, organic | ICD-9 | 327.0 |
| Insomnia, unspecified | ICD-9 | 780.52 |
| Insomnia, persistent / idiopathic | ICD-9 | 307.42 |
| Insomnia, transient | ICD-9 | 307.41 |
| Smell / taste dysfunction |  |  |
|  | ICD-10 | R43 |
|  | ICD-9 | 781.1 |
| Constipation |  |  |
|  | ICD-10 | K59.0 |
|  | ICD-9 | 564.0 |
| Urinary dysfunction |  |  |
|  | ICD-10 | R30-R39 |
|  | ICD-9 | 788 |
| Erectile dysfunction |  |  |
|  | ICD-10 | N52.(0,1,8,9] |
|  | ICD-9 | 607.84 |
| Orthostatic hypotension |  |  |
|  | ICD-10 | I95.1 |
|  | ICD-9 | 458.0 |

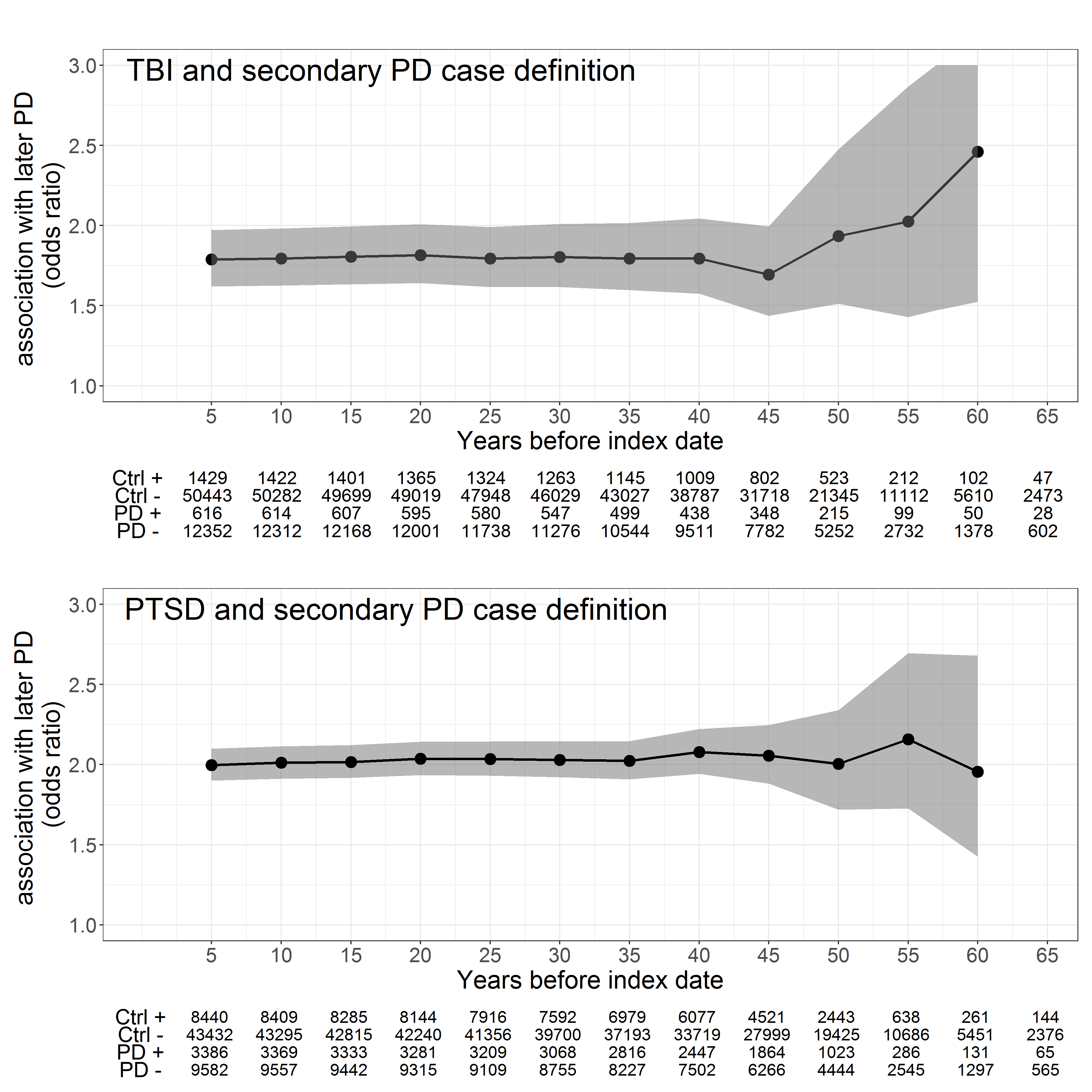

Supplement Figure 1: Association of Traumatic brain injury (TBI) and Post-traumatic stress disorder (PTSD) with the secondary VA neurology-based PD case definition. Odds ratios (black circles) and 95% confidence intervals (gray) shown at five-year intervals beginning at midpoint of active military duty and extending up to 5 years before the index date (year 0, date of PD diagnosis). Tables show number of cases and controls with (+) and without (-) trauma-related disorders.

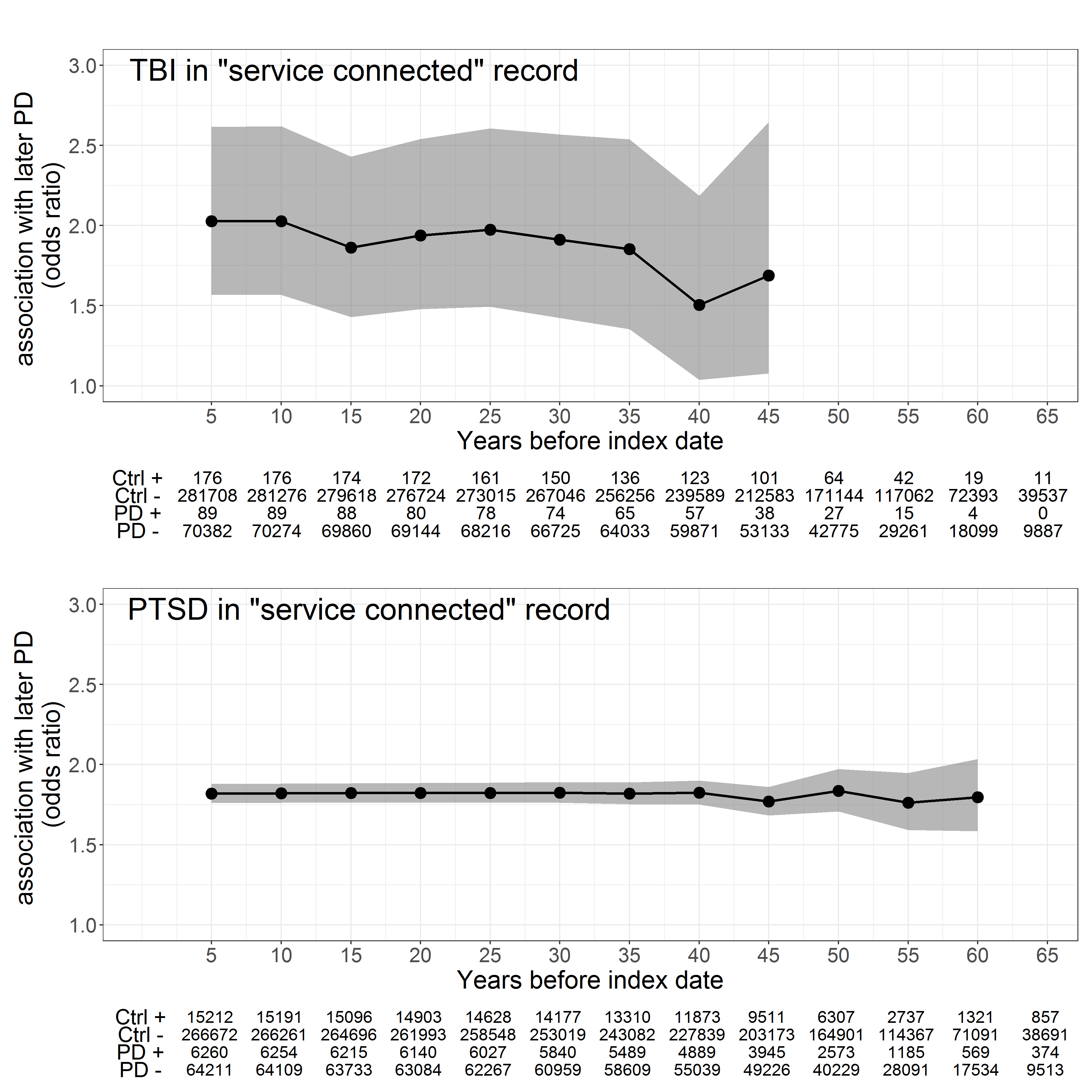

Supplement Figure 2 Association of PD in trauma-related disorders that are recorded with a positive flag in the medical records for being connected to active military service. Odds ratios (black circles) and 95% confidence intervals (gray) are shown for traumatic brain injury (TBI) and post-traumatic stress disorder (PTSD) at five-year intervals beginning at midpoint of active military duty and extending up to 5 years before the index date (year 0, date of PD diagnosis). Tables show number of cases and controls with (+) and without (-) trauma-related disorders.
